# Hand Hygiene Products and Adverse Skin Reactions: A Comparison between Health Care and Non-Health Care Workers

**DOI:** 10.1101/2022.02.15.22269519

**Authors:** Simanta Roy, Aishik Dipta Saha, Mohammad Azmain Iktidar, Sreshtha Chowdhury, Syed Md. Sayeem Tanvir, Mohammad Delwer Hossain Hawlader

**Affiliations:** Department of Public Health, North South University, Dhaka 1229, Bangladesh; Chattogram Medical College, 57 K.B. Fazlul Kader Rd, Chattogram 4203, Bangladesh

**Keywords:** Hand wash, Hand Hygiene, Skin problem, Skin Reaction, Sanitizer

## Abstract

**Background:** Since December 2019, a deadly coronavirus epidemic has swept the globe. Due to the significant risk of infection, frontline health workers had to use Personal Protective Equipment, including hand hygiene products, to keep their hands hygienic. The present study aims to compare adverse skin responses between Health Care Worker (HCW) and Non-Health Care Worker (NHCW).

**Materials and Methods:** A descriptive, cross-sectional study of HCW and NHCW throughout the country was executed. A self-structured questionnaire was utilized to gather data from 404 HCWs and 826 NHCWs during a two-month period using multistage sampling. STATA (v16) was used to analyse the data.

**Results:** 41.87% of the study participants experienced adverse skin reactions, which were more prevalent among HCW (65.10%) than NHCW (30.51%). The most often reported skin condition was skin dryness (34.39%), followed by skin peeling (11.71%). Users of alcohol-based hand sanitizers (ABHS) were more likely to get itch (8.13%), whereas soap water users were more likely to suffer skin peeling (35.74%) and rash (7.46%). There was a significant (p<0.001) association between occupation and adverse skin responses, with HCW being 3.5 times more likely to have adverse skin manifestations than NHCW.

**Conclusion:** The research showed that health care workers had a greater prevalence of skin conditions than the overall population. Hand hygiene guidelines for frontline employees should be equipped with instructions on how to protect oneself from these adverse skin manifestations, since frequent use constitutes a significant risk factor. Above all, health care professionals and the general population should be educated on good hand hygiene practices.

## Introduction

COVID-19 was declared a global pandemic by the World Health Organization on March 11, 2020, indicating a major global spread of an infectious illness.^1^ By then, the outbreak, which had initially begun in China in January 2020, had spread to 110 different countries, infecting over 118,000 people.^1^ Despite intensive public health precautions, the total number of confirmed cases worldwide has climbed to 250,154,972 as of this writing.^2^ By this time, the disease had also claimed 5,054,267 lives.^2^ As the disease spread, governments began to recognize its toll on the economy and health care sector. Hence, accelerated public health campaigns were put into motion in all forms of mass media. Schools were closed, travel bans were imposed, public gatherings were prohibited, and lockdowns were implemented. The World Health Organization laid out a guideline that enlisted a set of preventive measures to curb the exponential spread of COVID-19. Social distancing, wearing masks, immaculate hand hygiene, and vaccinations formed the pillars of the prevention strategy.

Among infection control tools, following face masks, hand hygiene is of utmost importance as hands might get contaminated directly via patients’ respiratory droplets from coughs and sneezes or indirectly via touching infected fomites. The CDC recommends hand hygiene by either using soap and water or ABHS.^3^ If soap and water are not available, using a hand sanitizer with a final concentration of at least 60% ethanol or 70% isopropyl alcohol inactivates SARS-CoV-2.^3^ Although the WHO advised using a palmful of alcohol-based solution and using it for twenty to thirty seconds, the guidance regarding the frequency of handwashing was vague.

As more people, both health care workers and non-healthcare professionals, began to adopt frequent hand washing with soap and water or ABHS, a series of adverse effects began to emerge, which appeared to be related to the use of these cleansing agents. This study takes a deep dive to explore these adverse effects, some of which have newly emerged and some of which were pre-existing conditions that appeared to have exacerbated. The aim was to establish an association between these effects and extensive hand hygiene and suggest appropriate changes to prevent or reduce these effects. The study also intended to compare the incidence of these events between health-care workers and the general population to further solidify the relationship between the determinants of hand hygiene products’ associated adverse effects.

## Materials and Methods

### Study participants and study site

This cross-sectional study surveyed 1230 respondents from Bangladesh’s eight divisions in a period of two months.

Initially, one government hospital, one private hospital, and one dental clinic were picked at random from a list of hospitals and dental clinics in each of the eight divisions (Dhaka, Chittagong, Rajshahi, Barisal, Mymensingh, Rangpur, Khulna, and Sylhet). A similar list was used to pick a military health-care institution. If any of the facilities chosen did not verbally concur, this selection approach was reconsidered. Later, using the snowball sampling approach, 404 health care personnel were chosen from these institutions.

Secondly, 826 consenting general citizens (non-health care professionals) were interviewed at public locations across the eight divisions using a fixed-step approach on a random route sample (every fifth person). To establish a representative sample of the general population per city, a quota sampling approach based on gender was applied (using data from the Bangladesh Bureau of Statistics). If a male subject is needed to fulfill the quota, every fifth person is contacted until a male is found.

### Data collection

Volunteers were recruited and trained at each of the research locations. After receiving the verbal agreement, volunteers conducted face-to-face interviews with subjects. A telephone interview was considered for health care personnel on COVID duty or who were isolated at home. Health care professionals included registered physicians, nurses, technicians, and assistants whereas non-health care workers were included in the general population. Due to the genetic and cultural diversity, foreign nationals were excluded.

### Study instrument

The researchers developed an initial questionnaire based on their review of existing studies. It was then pretested on 30 respondents, and the final version was accepted based on their recommendations. The questionnaire was translated from English to Bengali following ISPOR guideline.^4^ The final questionnaire retrieved information regarding respondents’ sociodemographic characteristics (age, sex, occupation, place of employment, work hours), hand hygiene product type, and frequency of usage (1-4, 5-10, 11-15, 16-20, >20). Additionally, the questionnaire included questions about the history of COVID-19 infection and vaccinations. Moreover, respondents were asked about any new skin problems associated with the usage of hand hygiene products. The researchers compiled a list of frequent and relevant dermatological problems from the responses. All the reporting was done according to Strengthening the Reporting of Observational Studies in Epidemiology (STROBE) guidelines.^5^

### Ethical consideration

The Institutional Review Board of North South University approved (Approval no 2021/OR-NSU/IRB/1001) the research protocol, which adhered to the 1975 Declaration of Helsinki’s ethical criteria (6th edition, 2008), as shown by a priori approval by the institutional review committee.

### Statistical analysis

The data has been processed and analysed using the Stata version 16.0. Categorical data were summarized using frequency and relative frequency and chi-square (χ^2^) test was used to explore the association of various adverse skin reactions to hand hygiene products. Finally, binary logistic regression model was fit to investigate the factors associated with adverse skin reactions among hand hygiene product users. Statistical significance level was set at p-value <0.05 and 95% confidence interval.

## Results

### Baseline characteristics of study participants

Table 1 depicts the background characteristics of the study population. Out of the entire study cohort, 826 (67.17%) were non-health care workers (NHCW) & 404 (32.83%) were health care workers (HCW). 56.83% of the respondents were male and 51.79% were literate up to graduate level. Most of the NHCW worked less than 8 hours a week (35.06%) but in the other hand, the majority of the HCW worked greater than 56 hours a week (24.26%). 75.53% of the entire study cohort used ABHS, while 69.35% used soap water. When asked about daily use frequency, 30.81% of the cohort reported that they had cleansed their hands 1 to 4 times a day, while 35.01% reported that they had used hand hygiene products 5 to 10 times a day. 19.95% of the HCW cleansed their hands greater than 20 times a day, while only 4.68% of the NHCW did the same. 31.22% of the entire cohort had a prior history of COVID-19 infection. Among HCW, 65.84% were infected and among NHCW, only 14.29% were infected. 60.89% of the study cohort was fully vaccinated.

**Table 1:**
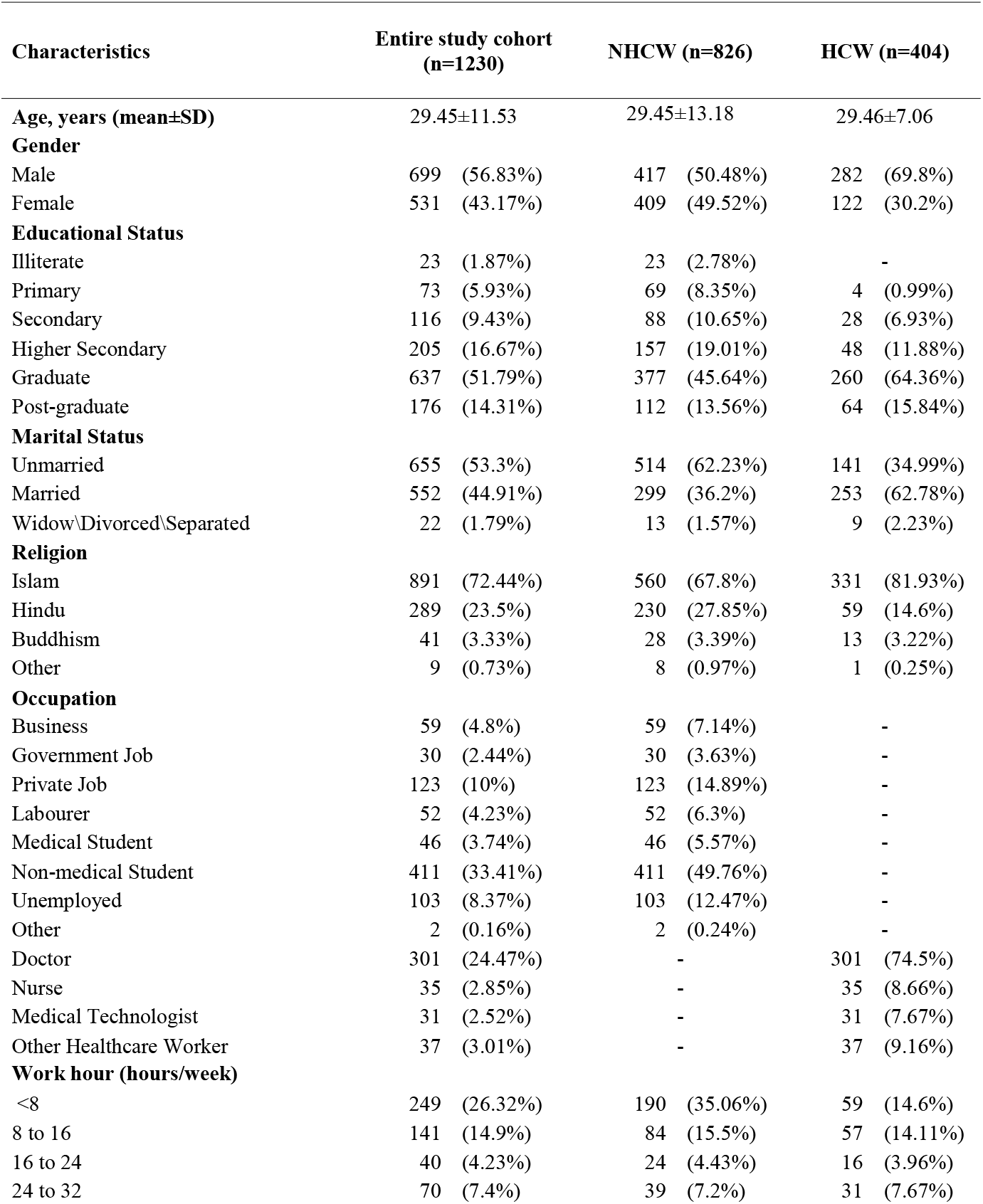
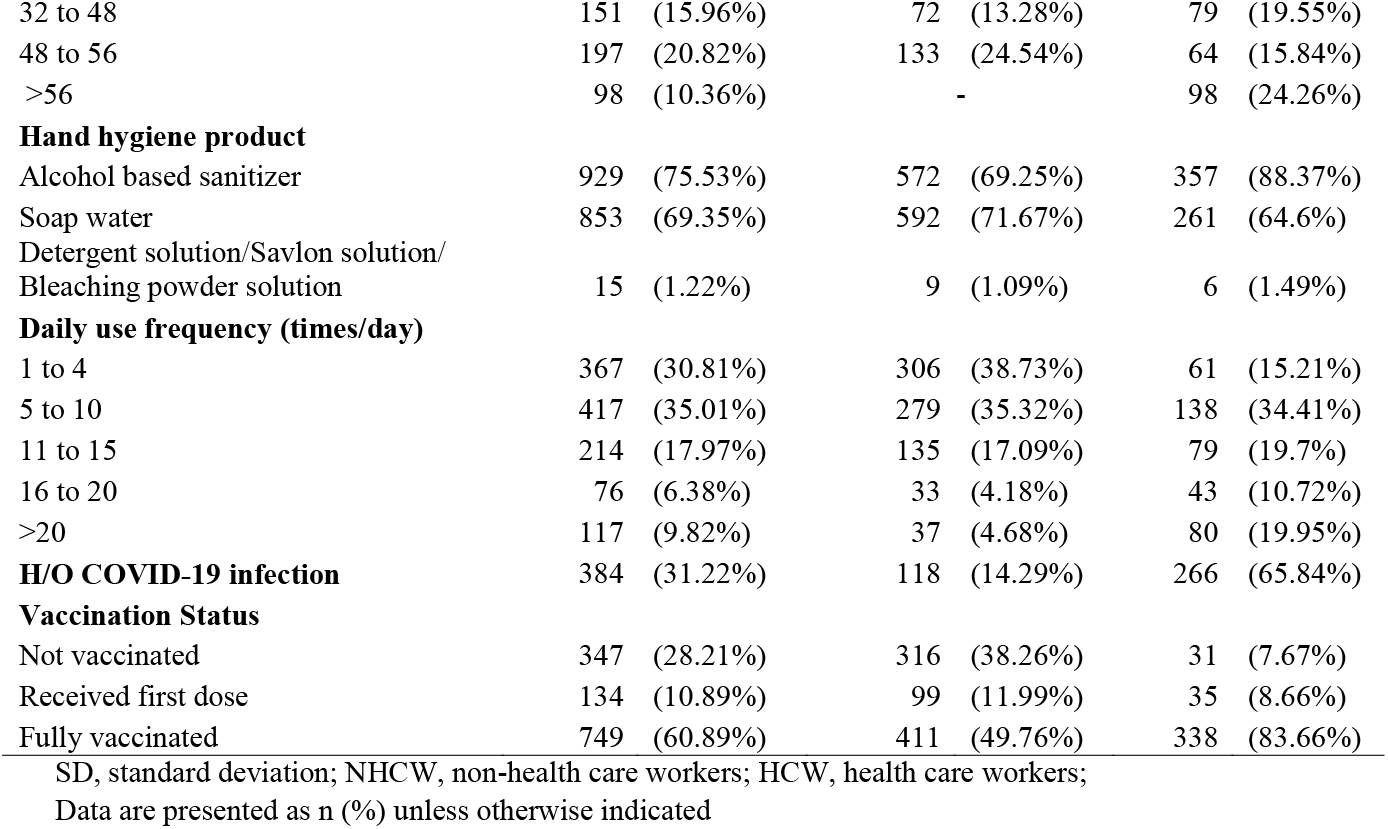
Background Characteristics of Study Population.

| Characteristics | Entire study cohort<br>(n=1230) |  | NHCW (n=826) |  | HCW (n=404) |  |
| --- | --- | --- | --- | --- | --- | --- |
| **Age, years (mean±SD)** | 29.45±11.53 |  | 29.45±13.18 |  | 29.46±7.06 |  |
| **Gender** |  |  |  |  |  |  |
| Male | 699 | (56.83%) | 417 | (50.48%) | 282 | (69.8%) |
| Female | 531 | (43.17%) | 409 | (49.52%) | 122 | (30.2%) |
| **Educational Status** |  |  |  |  |  |  |
| Illiterate | 23 | (1.87%) | 23 | (2.78%) | - |  |
| Primary | 73 | (5.93%) | 69 | (8.35%) | 4 | (0.99%) |
| Secondary | 116 | (9.43%) | 88 | (10.65%) | 28 | (6.93%) |
| Higher Secondary | 205 | (16.67%) | 157 | (19.01%) | 48 | (11.88%) |
| Graduate | 637 | (51.79%) | 377 | (45.64%) | 260 | (64.36%) |
| Post-graduate | 176 | (14.31%) | 112 | (13.56%) | 64 | (15.84%) |
| **Marital Status** |  |  |  |  |  |  |
| Unmarried | 655 | (53.3%) | 514 | (62.23%) | 141 | (34.99%) |
| Married | 552 | (44.91%) | 299 | (36.2%) | 253 | (62.78%) |
| Widow\Divorced\Separated | 22 | (1.79%) | 13 | (1.57%) | 9 | (2.23%) |
| **Religion** |  |  |  |  |  |  |
| Islam | 891 | (72.44%) | 560 | (67.8%) | 331 | (81.93%) |
| Hindu | 289 | (23.5%) | 230 | (27.85%) | 59 | (14.6%) |
| Buddhism | 41 | (3.33%) | 28 | (3.39%) | 13 | (3.22%) |
| Other | 9 | (0.73%) | 8 | (0.97%) | 1 | (0.25%) |
| **Occupation** |  |  |  |  |  |  |
| Business | 59 | (4.8%) | 59 | (7.14%) | - |  |
| Government Job | 30 | (2.44%) | 30 | (3.63%) | - |  |
| Private Job | 123 | (10%) | 123 | (14.89%) | - |  |
| Labourer | 52 | (4.23%) | 52 | (6.3%) | - |  |
| Medical Student | 46 | (3.74%) | 46 | (5.57%) | - |  |
| Non-medical Student | 411 | (33.41%) | 411 | (49.76%) | - |  |
| Unemployed | 103 | (8.37%) | 103 | (12.47%) | - |  |
| Other | 2 | (0.16%) | 2 | (0.24%) | - |  |
| Doctor | 301 | (24.47%) | - |  | 301 | (74.5%) |
| Nurse | 35 | (2.85%) | - |  | 35 | (8.66%) |
| Medical Technologist | 31 | (2.52%) | - |  | 31 | (7.67%) |
| Other Healthcare Worker | 37 | (3.01%) | - |  | 37 | (9.16%) |
| **Work hour (hours/week)** |  |  |  |  |  |  |
| <8 | 249 | (26.32%) | 190 | (35.06%) | 59 | (14.6%) |
| 8 to 16 | 141 | (14.9%) | 84 | (15.5%) | 57 | (14.11%) |
| 16 to 24 | 40 | (4.23%) | 24 | (4.43%) | 16 | (3.96%) |
| 24 to 32 | 70 | (7.4%) | 39 | (7.2%) | 31 | (7.67%) |

|  |  |  |  |
| --- | --- | --- | --- |
| 32 to 48 | 151 (15.96%) | 72 (13.28%) | 79 (19.55%) |
| 48 to 56 | 197 (20.82%) | 133 (24.54%) | 64 (15.84%) |
| >56 | 98 (10.36%) | - | 98 (24.26%) |
| **Hand hygiene product** |  |  |  |
| Alcohol based sanitizer | 929 (75.53%) | 572 (69.25%) | 357 (88.37%) |
| Soap water | 853 (69.35%) | 592 (71.67%) | 261 (64.6%) |
| Detergent solution/Savlon solution/<br>Bleaching powder solution | 15 (1.22%) | 9 (1.09%) | 6 (1.49%) |
| **Daily use frequency (times/day)** |  |  |  |
| 1 to 4 | 367 (30.81%) | 306 (38.73%) | 61 (15.21%) |
| 5 to 10 | 417 (35.01%) | 279 (35.32%) | 138 (34.41%) |
| 11 to 15 | 214 (17.97%) | 135 (17.09%) | 79 (19.7%) |
| 16 to 20 | 76 (6.38%) | 33 (4.18%) | 43 (10.72%) |
| >20 | 117 (9.82%) | 37 (4.68%) | 80 (19.95%) |
| **H/O COVID-19 infection** | 384 (31.22%) | 118 (14.29%) | 266 (65.84%) |
| **Vaccination Status** |  |  |  |
| Not vaccinated | 347 (28.21%) | 316 (38.26%) | 31 (7.67%) |
| Received first dose | 134 (10.89%) | 99 (11.99%) | 35 (8.66%) |
| Fully vaccinated | 749 (60.89%) | 411 (49.76%) | 338 (83.66%) |

SD, standard deviation; NHCW, non-health care workers; HCW, health care workers;
Data are presented as n (%) unless otherwise indicated

### Types of adverse skin reactions

Figure 2 depicts the types of adverse skin reactions and their frequencies following hand hygiene product use among the study population. Over half of the cohort reported no adverse reactions (56.1%), among which the majority were NHCW (67.55%). Adverse skin reactions were reported by 41.87% of the study population, which were more common among HCW (65.10%) compared to NHCW (30.51%). Skin dryness was the commonest reported skin problem (34.39%) followed by skin peeling (11.71%).

**Figure 1:**
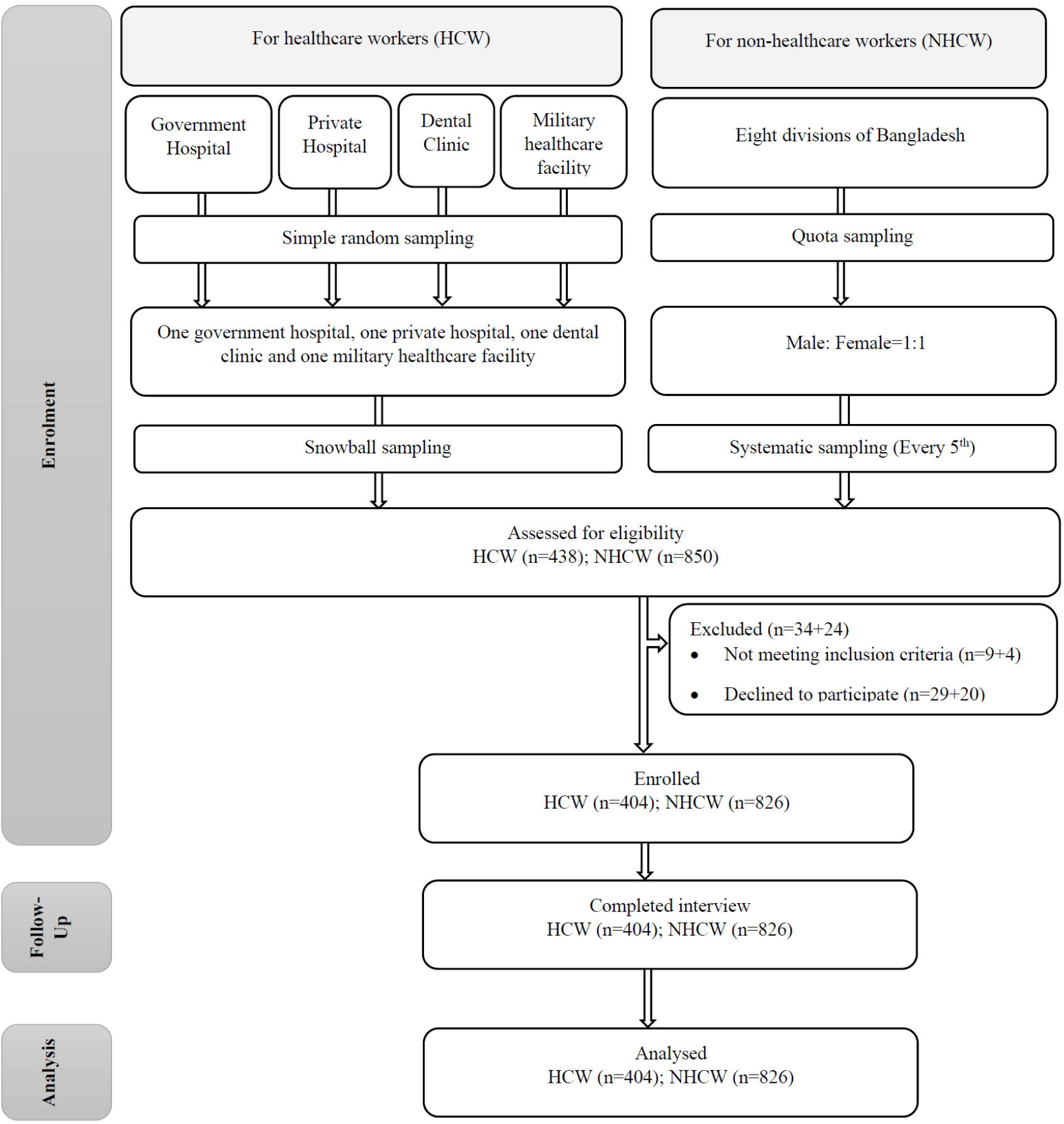

**Figure 2:**
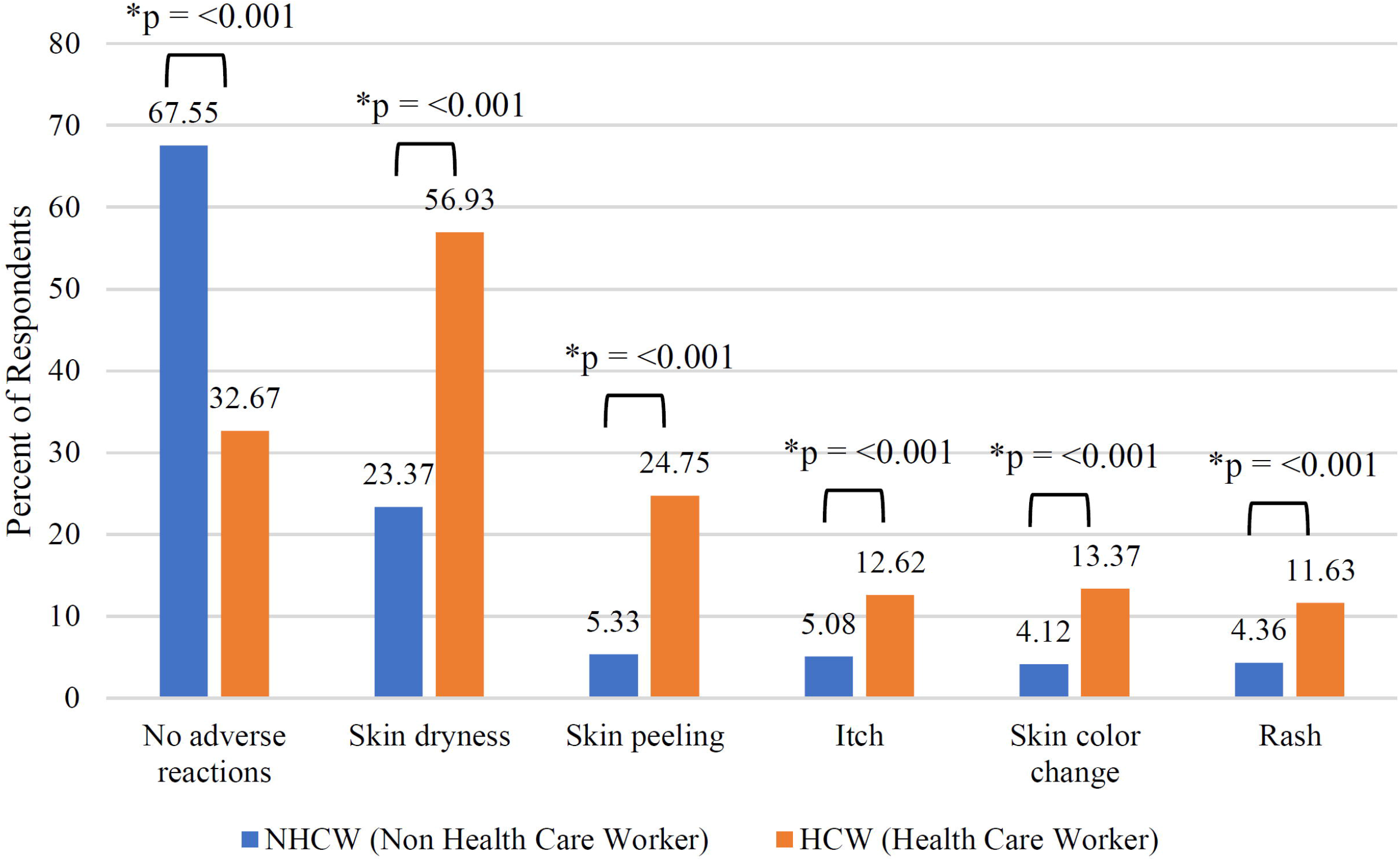

### Hand hygiene product use and adverse skin reactions

Figure 3 reveals study participants using soap water were more protected from developing adverse reactions (55.74%) compared to those using ABHS (53.01%) and detergent/savlon solution/bleaching powder solution (33.33%). Detergent/savlon solution/bleaching powder solution users were more prone to developing skin dryness (46.67%) and skin colour change (13.33%). On the other hand, ABHS users were more likely to develop itch (8.13%), and soap water users were susceptible to skin peeling (35.74%) and rash (7.46%).

**Figure 3:**
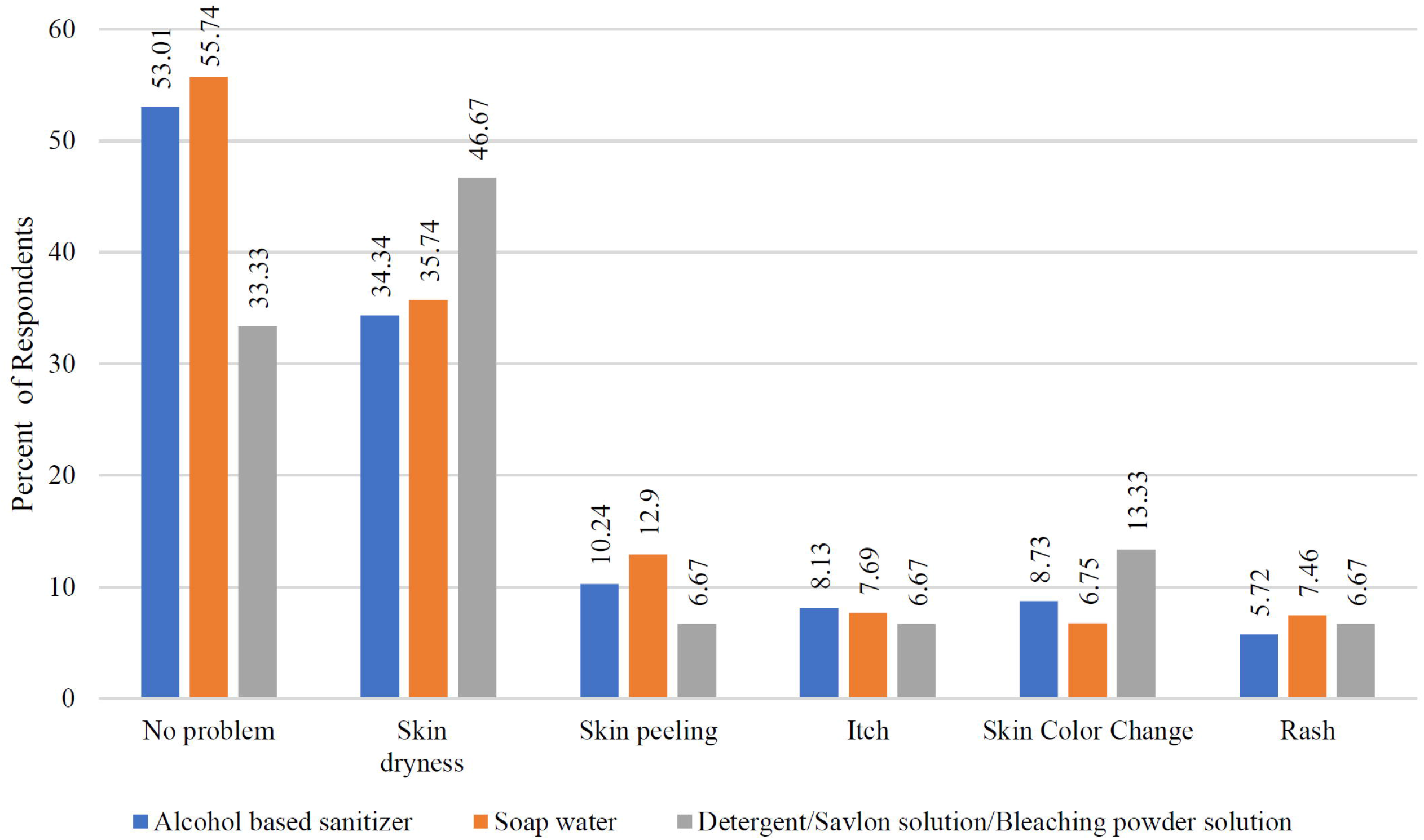

### Factors associated with adverse skin reactions

Unadjusted and adjusted results of binary logistic regression of the study variables are presented in **Table 2**. Age had a significant association with adverse skin reactions during both bivariate and multivariate analysis. Females were 1.92 times more likely to have adverse skin reactions compared to men. No significant association was found between adverse skin reaction and educational status. Regarding occupation, HCW were 3.5 times more likely to suffer from adverse skin manifestations that NHCW.

**Table 2:**
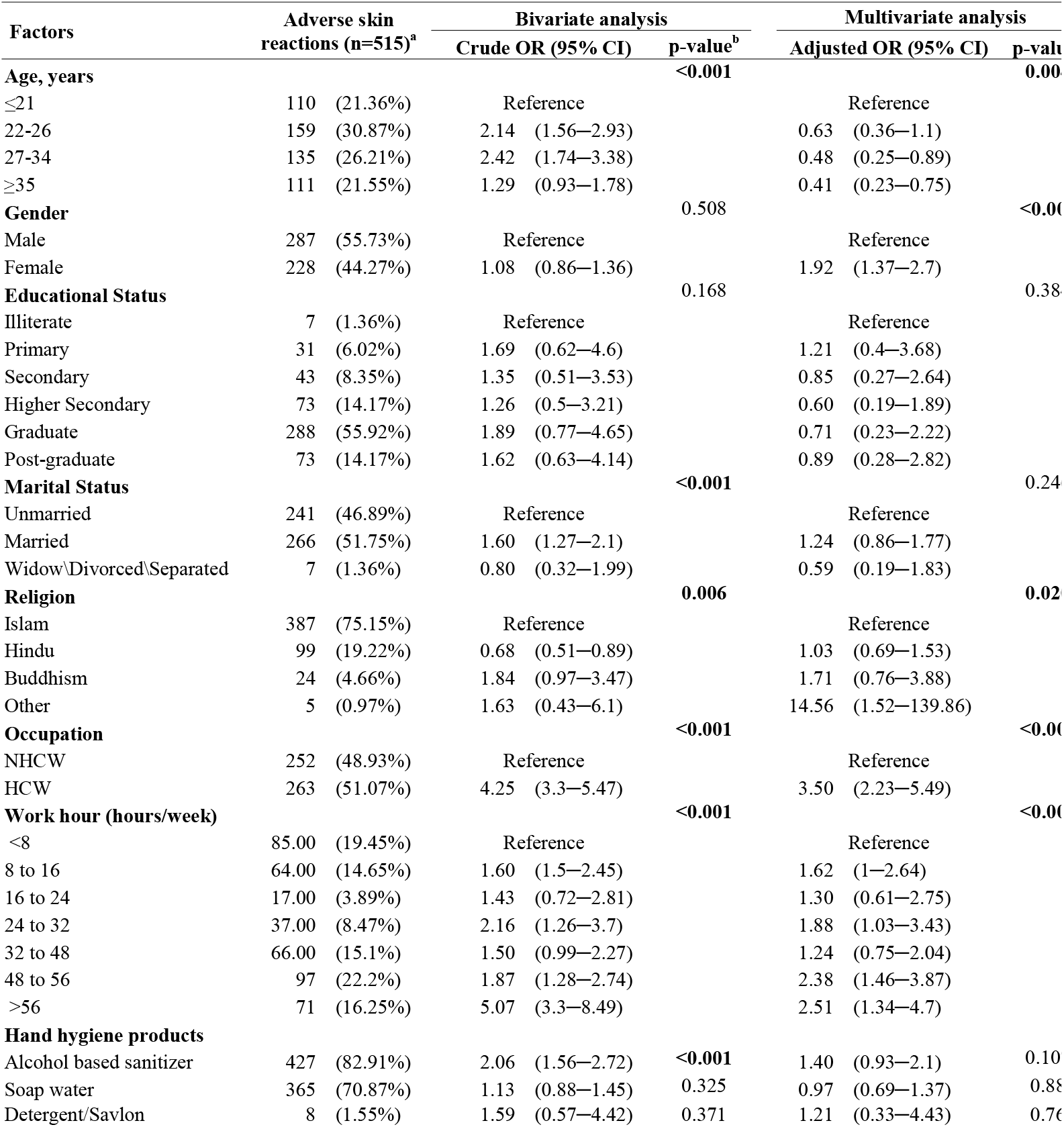
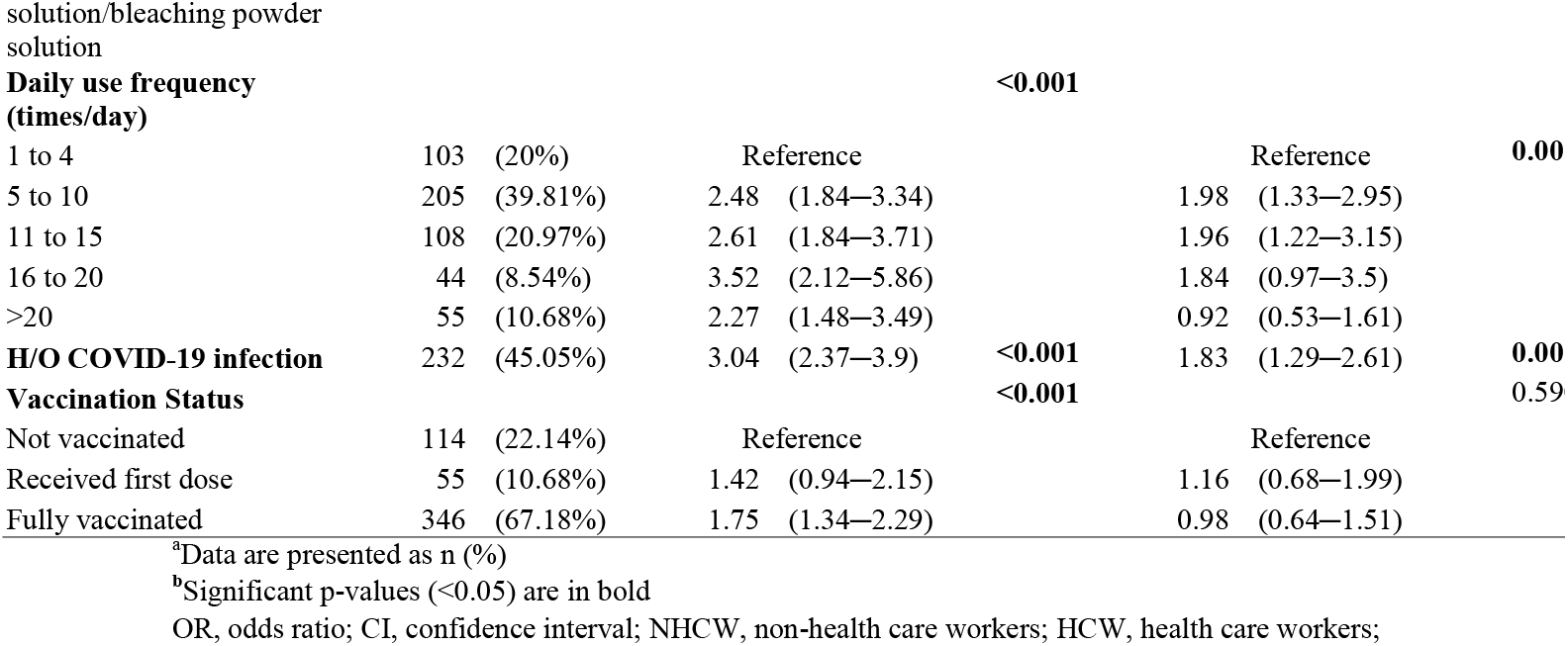
Factors associated with adverse skin reactions among the hand sanitizer users.

A multivariate analysis showed a significant association between work hours (hours/week) and adverse skin reactions. When compared to the group that worked fewer than 8 hours per week, those who worked between 48 and 56 hours and those who worked more than 56 hours were 2.38 times and 2.51 times more likely to experience adverse skin reactions, respectively. ABHS use was significantly associated with developing adverse reactions in bivariate analysis (Crude OR: 2.06; 95% CI: 1.56-2.72. p<0.001). Although it loses significance in the multivariate analysis, ABHS users were 40% more likely to develop adverse skin reactions, whereas soap water users were 3% less likely. The daily use frequency of hand hygiene products had a significant (p<0.001) association with adverse skin manifestations. The group that used ABHS 5 to 10 times per day had almost two times the risk of having adverse skin responses compared to the group that used ABHS 1 to 4 times per day. Subjects who had a history of COVID-19 infection were nearly two times more likely to develop adverse skin reactions than those who did not (p = 0.001). No significant association was found between vaccination status and adverse skin reactions.

## Discussion

In the study, adverse skin responses were reported by 41.87% of the study participants, with HCWs (65.10 %) reporting them more frequently than NHCWs (30.51 %). A highly significant association between occupation and adverse skin reactions has been revealed, where HCW were 3.5 times more likely to suffer from adverse skin manifestations than NHCW. This could be due to the higher frequency of use of hand hygiene products as well as longer working hours of HCW compared to NHCW. This has been supported by a multiple logistic regression model, which showed a significant association between work hours (hours/week) and adverse skin reaction. Moreover, health-care professionals must maintain strict hand hygiene and wear gloves for lengthy periods. For them, a moisturizer under gloves can also be effective. Moisturizers with a water base are safe under all gloves; however, oil-based moisturizers can break down latex and rubber by making the material swell or become brittle.^6^

Dermatological problems have been linked to frequent handwashing even before the current outbreak. Following strict hygiene practices during the 2014 Ebola epidemic, health-care workers developed hand eczema, which was worse when using soap than ABHS.^7^ According to our study, the daily use frequency of hand hygiene products had a significant association with adverse skin manifestations. When compared to those who used ABHS 1 to 4 times per day, those who used ABHS 5 to 10 times per day had a nearly double risk of developing unfavorable skin reactions. In China, 74.5% of health professionals had hand eczema, and handwashing frequency (>10 times/day) was a significant predictor.^8^ In Germany, handwashing frequency-doubled, resulting in a 90.2% incidence of skin problems.^9^ India and Italy have reported increasing teledermatology consultations for hand eczema.^10,11^ An Indian research identified 16 cases of hand eczema in 10 days due to frequent hand sanitizer use/washing.^10^ Therefore, stringent hand cleanliness methods put both the general population and health professionals at risk for skin-related issues. So, it may be prudent to limit hand hygiene products to a certain time to reduce the risk of adverse skin reactions. Limiting the frequency may reduce the incidence of adverse skin reactions but might not always be practically feasible while adhering to strict infection control protocols.

Skin dryness was the commonest reported skin problem (34.39%), followed by skin peeling (11.71%). ABHS have a low irritating potential and are more tolerable, but they tend to make the skin dry and more susceptible to contact dermatitis.^12^ To alleviate this most commonly encountered adverse skin reaction, both the WHO and the American Contact Dermatitis Society have advocated the use of moisturizers after hand cleansing.^6,13^ However, it has been advised to avoid moisturizers in jars to prevent double dipping into and potentially to contaminate the product. Instead, moisturizers in pocket-sized tubes were suggested to keep in one’s person for frequent reapplication.

In our study, participants using soap water were more protected from developing adverse reactions (55.74%) compared to those using ABHS (53.01%) and detergent/savlon solution/bleaching powder solution (33.33%). In general, while using ABHS, it is better to avoid ones with allergenic surfactants, preservatives, fragrances, or dyes.^6^ ABHS with added moisturizers has been shown to be beneficial in ameliorating hand dryness associated with frequent hand hygiene.^6^ Moreover, for the formulation, ethanol is preferable since it is less irritating than isopropyl alcohol despite equal effectiveness.^14^

When exploring the association between gender and adverse skin reactions, females were almost twice as likely to have adverse skin reactions compared to men. There was no statistically significant association between COVID-19 vaccination status and hand hygiene-related adverse skin conditions. Subjects who have had a history of COVID-19 infection were nearly two times more likely to suffer from adverse cutaneous manifestations. However, why such an association exists still owes an explanation and demands further studies to explore the matter. One of the few limitations of our study is that hand hygiene technique was not taken into consideration when finding out the prevalence of adverse skin reactions and its relation to hand hygiene product use. The WHO and American Contact Dermatitis Society recommend cleansing by gentle patting rather than by rubbing to minimize mechanical trauma to the skin barrier.^6,13^ Besides, the effect of gloves usage and temperature of water used for handwashing with soap was not considered while conducting the study.^6,15^ Applying gloves when hands are still wet from either hand washing or ABHS is also not recommended because the risk for skin irritation due to trapping of irritating ingredients increases.^16^

It’s essential to note some of our study’s limitations as well as the methods we used to solve them. Due to our study’s cross-sectional nature, we cannot infer causality for the associations that we have presented in this paper. However, by presenting AORs using multiple regression models, we attempted to account for the potential effect of confounders. All the problems presented in this study were self-reported by the respondents, which may have led to self-reporting bias. In order to minimize this, all our data collectors were medical students with adequate training and followed a predefined neutral script. We also acknowledge that the findings of our study may not be generalizable due to our sample size. In order to make the sample representative and reduce selection bias, the health care workers were selected by snowball sampling from randomly selected health care facilities, and non-healthcare workers were selected by quota sampling across all the divisions of Bangladesh.

## Conclusion

Our research found that first-line health care providers had a significantly higher prevalence of adverse skin reactions from hand hygiene product use than the overall population. Moreover, a higher frequency of hand hygiene product use was a major risk factor, highlighting the need for the inclusion of protective measures in these hand hygiene guidelines for frontline workers. Most importantly, both health workers and the general public should be educated on reasonable and sensible hand hygiene.

## Data Availability

All data produced in the present work are contained in the manuscript

